# Reconciling early-outbreak estimates of the basic reproductive number and its uncertainty: framework and applications to the novel coronavirus (SARS-CoV-2) outbreak

**DOI:** 10.1101/2020.01.30.20019877

**Authors:** Sang Woo Park, Benjamin M. Bolker, David Champredon, David J. D. Earn, Michael Li, Joshua S. Weitz, Bryan T. Grenfell, Jonathan Dushoff

## Abstract

A novel coronavirus (SARS-CoV-2) has recently emerged as a global threat. As the epidemic progresses, many disease modelers have focused on estimating the basic reproductive number *ℛ*_0_– the average number of secondary cases caused by a primary case in an otherwise susceptible population. The modeling approaches and resulting estimates of *ℛ*_0_ vary widely, despite relying on similar data sources. Here, we present a novel statistical framework for comparing and combining different estimates of *ℛ*_0_ across a wide range of models by decomposing the basic reproductive number into three key quantities: the exponential growth rate *r*, the mean generation interval 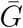, and the generation-interval dispersion *κ*. We then apply our framework to early estimates of *ℛ*_0_ for the SARS-CoV-2 outbreak. We show that many early *ℛ*_0_ estimates are overly confident. Our results emphasize the importance of propagating uncertainties in all components of *ℛ*_0_, including the shape of the generation-interval distribution, in efforts to estimate *ℛ*_0_ at the outset of an epidemic.

## 1 Introduction

Since December 2019, a novel coronavirus (SARS-CoV-2) has been spreading in China and other parts of the world (World Health Organization, 2020d). Although the virus is believed to have originated from animal reservoirs (Centers for Disease Control and Prevention, 2020), the ability of SARS-CoV-2 ability to directly transmit between humans has posed a greater threat for its spread (Huang et al., 2020; World Health Organization, 2020c). As of February 27, 2020, the World Health Organization (WHO) has confirmed 82,294 cases of the coronavirus disease (COVID-19), including 3,664 confirmed cases in 46 different countries, outside China (World Health Organization, 2020a).

As the disease continues to spread, many researchers have already published their analyses of the outbreak as pre-prints (e.g., Bedford et al. (2020); Imai et al. (2020); Liu et al. (2020); Majumder and Mandl (2020); Read et al. (2020a); Zhao et al. (2020)) and in peer-reviewed journals (e.g., Li et al. (2020); Riou and Althaus (2020b); Wu et al. (2020); Zhao et al. (2020)), focusing in particular on estimates of the basic reproductive number *ℛ*_0_ (i.e., the average number of secondary cases generated by a primary case in a fully susceptible population (Anderson and May, 1991; Diekmann et al., 1990)). Estimates of the basic reproductive number are of interest during an outbreak because they provide information about the level of intervention required to interrupt transmission (Anderson and May, 1991), and about the potential final size of the outbreak (Anderson and May, 1991; Ma and Earn, 2006). We commend these researchers for their timely contribution and those who made the data publicly available. However, it can be difficult to compare a disparate set of estimates of *ℛ*_0_ from different research groups (as well as the associated degrees of uncertainty) when the estimation methods and their underlying assumptions vary widely.

Here, we show that a wide range of approaches to estimating *ℛ*_0_ can be understood and compared in terms of estimates of three quantities: the exponential growth rate *r*, the mean generation interval 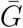, and the generation-interval dispersion *κ*. The generation interval, defined as the interval between the time when an individual becomes infected and the time when that individual infects another individual (Svensson, 2007), plays a key role in shaping the relationship between *r* and *ℛ*_0_ (Wearing et al., 2005; Roberts and Heesterbeek, 2007; Wallinga and Lipsitch, 2007; Park et al., 2019); therefore, estimates of *ℛ*_0_ from different models directly depend on their implicit assumptions about the generation-interval distribution and the exponential growth rate. Early in an epidemic, information is scarce and there is inevitably a great deal of uncertainty surrounding both case reports (affecting the estimates of the exponential growth rate) and contact tracing (affecting the estimates of the generation-interval distribution). We suggest that disease modelers should make sure their assumptions about these three quantities are clear and reasonable, and that estimates of uncertainty in *ℛ*_0_ should propagate error from all three sources (Elderd et al., 2006).

We compare seven disparate models published online between January 23–26, 2020 that estimated *ℛ*_0_ for the SARS-CoV-2 outbreak (Bedford et al., 2020; Imai et al., 2020; Liu et al., 2020; Majumder and Mandl, 2020; Read et al., 2020a; Riou and Althaus, 2020a; Zhao et al., 2020). We use a Bayesian multilevel model to construct pooled estimates for the three key quantities: *r*, 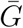, and *κ*; the pooled estimates reflect the uncertainties present in modeling approaches and their underlying assumptions. We use these pooled estimates to illustrate the importance of propagating different sources of error, particularly uncertainty in both the growth rate and the generation interval. We also use our framework to tease apart which assumptions of these different models led to their different estimates and confidence intervals. Despite the availability of more recent and/or updated estimates of *ℛ*_0_, we restrict ourselves to the estimates above in order to focus on the resolution of uncertainty in the earliest stages of an epidemic.

## 2 Methods

### 2.1 Description of the studies

We gathered information on estimates of *ℛ*_0_ and their assumptions about the underlying generation-interval distributions from 7 articles that were published online between January 23–26, 2020 (Table 1). Five studies (Liu et al., 2020; Majumder and Mandl, 2020; Read et al., 2020a; Riou and Althaus, 2020a; Zhao et al., 2020) were uploaded to pre-print servers (bioRxiv, medRxiv, and SSRN); one report was posted on the web site of Imperial College London (Imai et al., 2020); and one report was posted on nextstrain.org (Bedford et al., 2020). Their modeling approaches vary widely: a branching process model (Bedford et al., 2020; Imai et al., 2020; Riou and Althaus, 2020a), a deterministic Susceptible-Exposed-Infected-Recovered (SEIR) model (Read et al., 2020a), an exponential growth model (Zhao et al., 2020), a Poisson offspring distribution model (Liu et al., 2020), and the Incidence Decay and Exponential Adjustment (IDEA) model (Majumder and Mandl, 2020). Four studies estimated *ℛ*_0_ by directly fitting their models to incidence data (Read et al., 2020a; Zhao et al., 2020; Liu et al., 2020; Majumder and Mandl, 2020). The remaining three studies estimated *ℛ*_0_ by comparing the predicted number of cases from their models with the estimated number of total cases by January 18 (between 1,000 and 9,7000 (Imai et al., 2020)) Some of these studies have now been published in peer-reviewed journals (Riou and Althaus, 2020b; Zhao et al., 2020) or have been updated with better uncertainty quantification (Read et al., 2020b).

**Table 1:** Reported estimates of the basic reproductive number and the assumptions about the generation-interval distributions. Estimates of *ℛ*_0_ and their assumptions about the shape of the generation interval distributions were collected from 7 studies. ^*∗*^We treat these intervals as a 95% confidence interval in our analysis. ^*†*^We assume *κ* = 0.5 in our analysis. ^*‡*^The authors presented *ℛ*_0_ estimates under different assumptions regarding the reporting rate; we use their baseline scenario in our analysis to remain consistent with other studies, which do not account for changes in the reporting rate.

| | Basic reproductive number $\mathcal{R}_0$ | Mean generation interval $\bar{G}$ (days) | Generation-interval dispersion $\kappa$ | |
| --- | --- | --- | --- | --- |
| Study 1 | 1.5–3.5 | 10 | 1 | Bedford et al. (2020) |
| Study 2 | 2.5 (1.5–3.5)* | 8.4 | unspecified<sup>†</sup> | Imai et al. (2020) |
| Study 3 | 2.92 (95% CI: 2.28–3.67) | 8.4 | 0.2 | Liu et al. (2020) |
| Study 4 | 3.8 (95% CI: 3.6–4.0) | 7.6 | 0.5 | Read et al. (2020a) |
| Study 5 | 2.2 (90% CI: 1.4–3.8) | 7–14 | 0.5 | Riou and Althaus (2020a) |
| Study 6 | 5.47 (95% CI: 4.16–7.10)<sup>‡</sup> | 7.6–8.4 | 0.2 | Zhao et al. (2020) |
| Study 7 | 2.0–3.1 | 6–10 | 0 | Majumder and Mandl (2020) |

### 2.2 Gamma approximation framework for linking *r* and *R*_0_

Early in an outbreak, *ℛ*_0_ is difficult to estimate directly; instead, *ℛ*_0_ is often inferred from the exponential growth rate *r*, which can be estimated reliably from incidence data (Ma et al., 2014). Given an estimate of the exponential growth rate *r* and an *intrinsic* generation-interval distribution *g*(*τ*) (Champredon and Dushoff, 2015), the basic reproductive number can be estimated via the Euler-Lotka equation (Wallinga and Lipsitch, 2007):

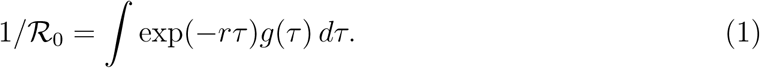

In other words, estimates of *ℛ*_0_ must depend on the assumptions about the exponential growth rate *r* and the shape of the generation-interval distribution *g*(*τ*).

Here, we use the gamma approximation framework (Park et al., 2019) to (i) characterize the amount of uncertainty present in the exponential growth rates and the shape of the generation-interval distribution and (ii) assess the degree to which these uncertainties affect the estimate of *ℛ*_0_. Assuming that generation intervals follow a gamma distribution with the mean 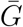 and the squared coefficient of variation *κ*, we have

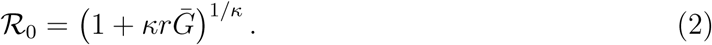

This equation demonstrates that a generation-interval distribution that has a larger mean (higher 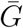) or is less variable (lower *κ*) will give a higher estimate of *ℛ*_0_ for the same value of *r* (Wallinga and Lipsitch, 2007).

### 2.3 Statistical framework

As most studies do not report their estimates of the exponential growth rate, we first recalculate the exponential growth rate that correspond to their model assumptions. We do so by modeling reported distributions of the reproductive number *ℛ*_0_, the mean generation interval, 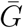 and the generation-interval dispersion parameter *κ* with appropriate probability distributions; we used gamma distributions to model values reported with confidence intervals and uniform distributions to model values reported with ranges. For example, Study 3 estimated *ℛ*_0_ = 2.92 (95% CI: 2.28–3.67); we model this estimate as a gamma distribution with a mean of 2.92 and a shape parameter of 67, which has a 95% probability of containing a value between 2.28 and 3.67 (see Table 2 for a complete description). For each study *i*, we construct a family of parameter sets by drawing 100,000 random samples from the probability distributions (Table 2) that represent the estimates of *ℛ*_0*i*_ and the assumed values of 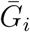 and *κ*_*i*_ and calculate the exponential growth rate *r*_*i*_ via the inverse of Eq. 2:

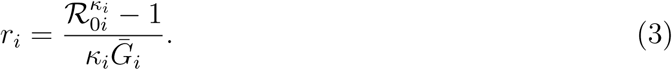

**Table 2:** Probability distributions for *ℛ*_0_, 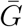, and *κ*. We use these probability distributions to obtain a probability distribution for the exponential growth rate *r*. The gamma distribution is parameterized by its mean and shape *α*. Constant values are fixed according to Table 1. ^*∗*^We do not account for this uncertainty during our recalculation of the exponential growth rate *r* because the reported estimate of *ℛ*_0_ and its uncertainty assumes 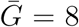; the original article reports three *ℛ*_0_ (and 95% CIs) estimates using three different values of 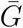: 7.6 (MERS-like), 8 (average), and 8.4 (SARS-like). We still account for this uncertainty in our pooled estimates (*µ*_*G*_). ^*†*^Study 6 uses the IDEA model (Fisman et al., 2013), through which the authors effectively fit an exponential curve to the cumulative number of confirmed cases without propagating any statistical uncertainty. Instead of modeling *ℛ*_0_ with a probability distribution and recalculating *r*, we use *r* = 0.114 days^*−*1^, which explains all uncertainty in the reported *ℛ*_0_, when combined with the considered range of 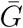.

| | Basic reproductive number $\mathcal{R}_0$ | Mean generation interval $\bar{G}$ (days) | Generation-interval dispersion $\kappa$ |
| --- | --- | --- | --- |
| Study 1 | Uniform(1.5, 3.5) | 10 | 1 |
| Study 2 | Gamma(mean = 2.6, $\alpha$ = 28) | 8.4 | 0.5 |
| Study 3 | Gamma(mean = 2.92, $\alpha$ = 67) | 8.4 | 0.2 |
| Study 4 | Gamma(mean = 3.8, $\alpha$ = 1400) | 7.6 | 0.5 |
| Study 5 | Gamma(mean = 2.2, $\alpha$ = 12) | Uniform(7, 14) | 0.5 |
| Study 6 | Gamma(mean = 5.47, $\alpha$ = 54) | Uniform(7.6, 8.4)* | 0.2 |
| Study 7 | exp($r\bar{G}$)<sup>†</sup> | Uniform(6, 10) | 0 |

This allows us to approximate the probability distributions of the estimated exponential growth rates by each study; uncertainties in the probability distributions that we calculate for the estimated exponential growth rates will reflect the methods and assumptions that the studies rely on.

We construct pooled estimates for each parameter (*r*, 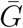, and *κ*) using a Bayesian multilevel modeling approach, which assumes that the parameters across different studies come from the same gamma distribution. The pooled estimates, which are represented as probability distributions rather than point estimates, allow us to average across different modeling approaches, while accounting for the uncertainties in the assumptions they make:

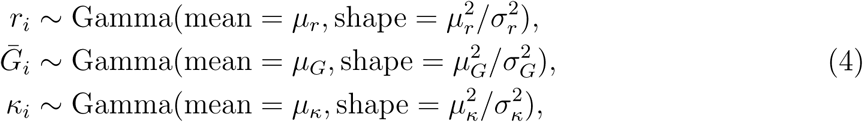

where *µ*_*r*_, *µ*_*G*_, *µ*_*κ*_ represent the pooled estimates, and *σ*_*r*_, *σ*_*G*_, and *σ*_*κ*_ represent between-study standard deviations. We account for uncertainties associated with *r*_*i*_, 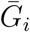 and *κ*_*i*_ (and their correlations), by drawing a random set from the family of parameter sets for each study at each Metropolis-Hastings step. Since the gamma distribution does not allow zeros, we use *κ* = 0.02 instead for Study 7. We note that this approach does not account for nonindependence between the parameter estimates made by different modelers. As we add more models, the pooled estimates can become sharper even when the models no longer add more information. Thus, the pooled estimator should be interpreted with care.

We use weakly informative priors on hyperparameters:

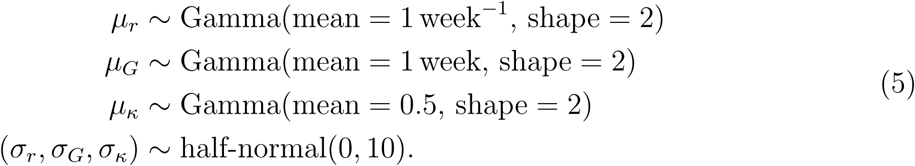

We followed recommendations outlined in Gelman et al. (2006), parameterizing the top-level gamma distributions in terms of their means and standard deviations and imposing weakly informative prior distributions on between-study standard deviations, i.e., half-normal(0, 10). We had initially used gamma priors with small shape parameters (*<* 1) on between-study shape parameters (= *µ*^2^*/σ*^2^) but found this put too much prior probability on large between-study variances. This phenomenon is a known problem (Gelman et al., 2006). Alternative choices of prior for the between-study shape parameters are also suboptimal: imposing strong priors (e.g. half-*t*(*µ* = 0, *σ* = 1, *ν* = 4) assumes *a priori* that between-study variance is large, while weak priors (e.g. half-Cauchy(0,5)) can lead to poor mixing.

We run 4 independent Markov Chain Monte Carlo chains each consisting of 500,000 burnin steps and 500,000 sampling steps. Posterior samples are thinned every 1000 steps. Convergence is assessed by ensuring that the Gelman-Rubin statistic is below 1.01 for all hyperparameters (Gelman et al., 1992); trace plots and marginal posterior distribution plots are presented in Appendix. 95% confidence intervals are calculated by taking 2.5% and 97.5% quantiles from the marginal posterior distribution for each parameter.

## 3 Results

Fig. 1 compares the reported values of the exponential growth rate *r*, mean generation interval 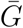, and the generation-interval dispersion *κ* from different studies with the pooled estimates that we calculate from our multilevel model. We find that there is a large uncertainty associated with the underlying parameters; many models rely on stronger assumptions that ignore these uncertainties. Surprisingly, no studies take into account how the variation in generation intervals affects their estimates of *ℛ*_0_: all studies assumed fixed values for *κ*, ranging from 0 to 1. Assuming fixed parameter values can lead to overly strong conclusions (Elderd et al., 2006).

**Figure 1:**
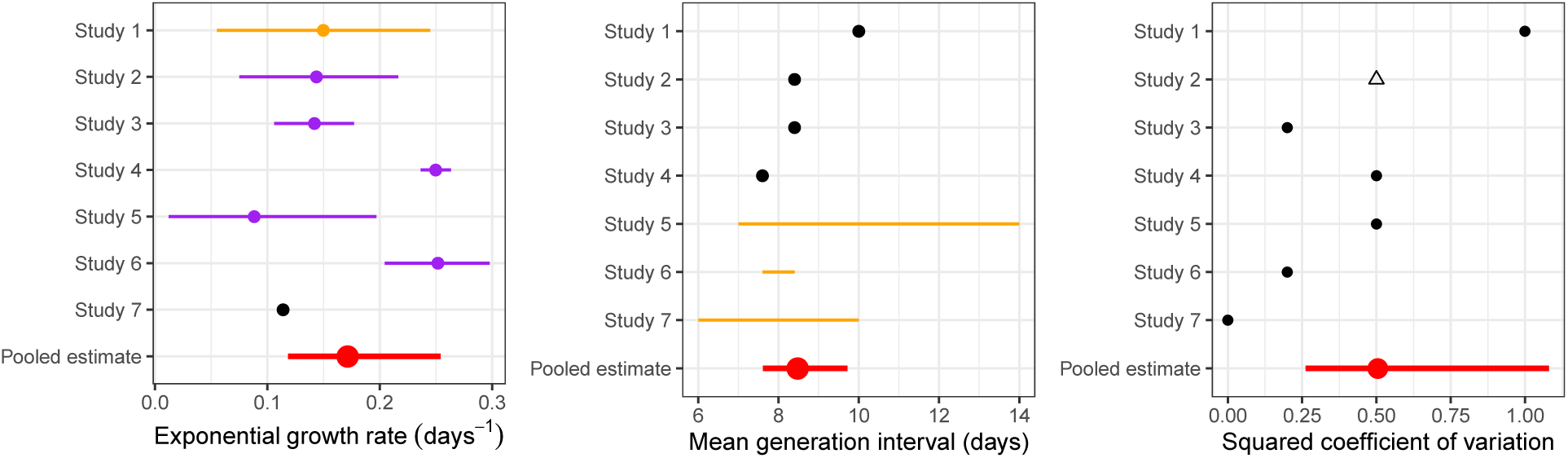
Comparisons of the reported parameter values with our pooled estimate. We inferred point estimates (black), uniform distributions (orange) or confidence intervals (purple) for each parameter from each study, and combined them into pooled estimates (red; see text). Open triangle: we assumed *κ* = 0.5 for Study 2 which does not report generation-interval dispersion.

Fig. 2 shows how propagating uncertainty in different combinations would affect estimates and CIs for *ℛ*_0_. For illustrative purposes, we use our pooled estimates, which may represent a reasonable proxy for the state of knowledge as of January 23–26 (Fig. 2A). Comparing the models that include only some sources of uncertainty to the “all” model, we see that propagating error from the growth rate (which all but one of the studies reviewed did) is absolutely crucial: the middle bar (“GI mean”), which lacks growth-rate uncertainty, is relatively narrow. In this case, propagating error from the mean generation interval has negligible effect compared to propagating the uncertainty in *r*. Uncertainty in the generation-interval dispersion also has important effects as it determines the functional form of the relationship between *r* and *ℛ*_0_ (compare “growth rate + GI mean” with “all”). For example, reducing the dispersion parameter *κ* from 1 (assuming exponentially distributed generation intervals) to 0 (assuming fixed generation intervals) changes the *r*– *ℛ*_0_ relationship from linear to exponential, therefore increasing the sensitivity of *ℛ*_0_ estimates to *r* and 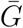.

**Figure 2:**
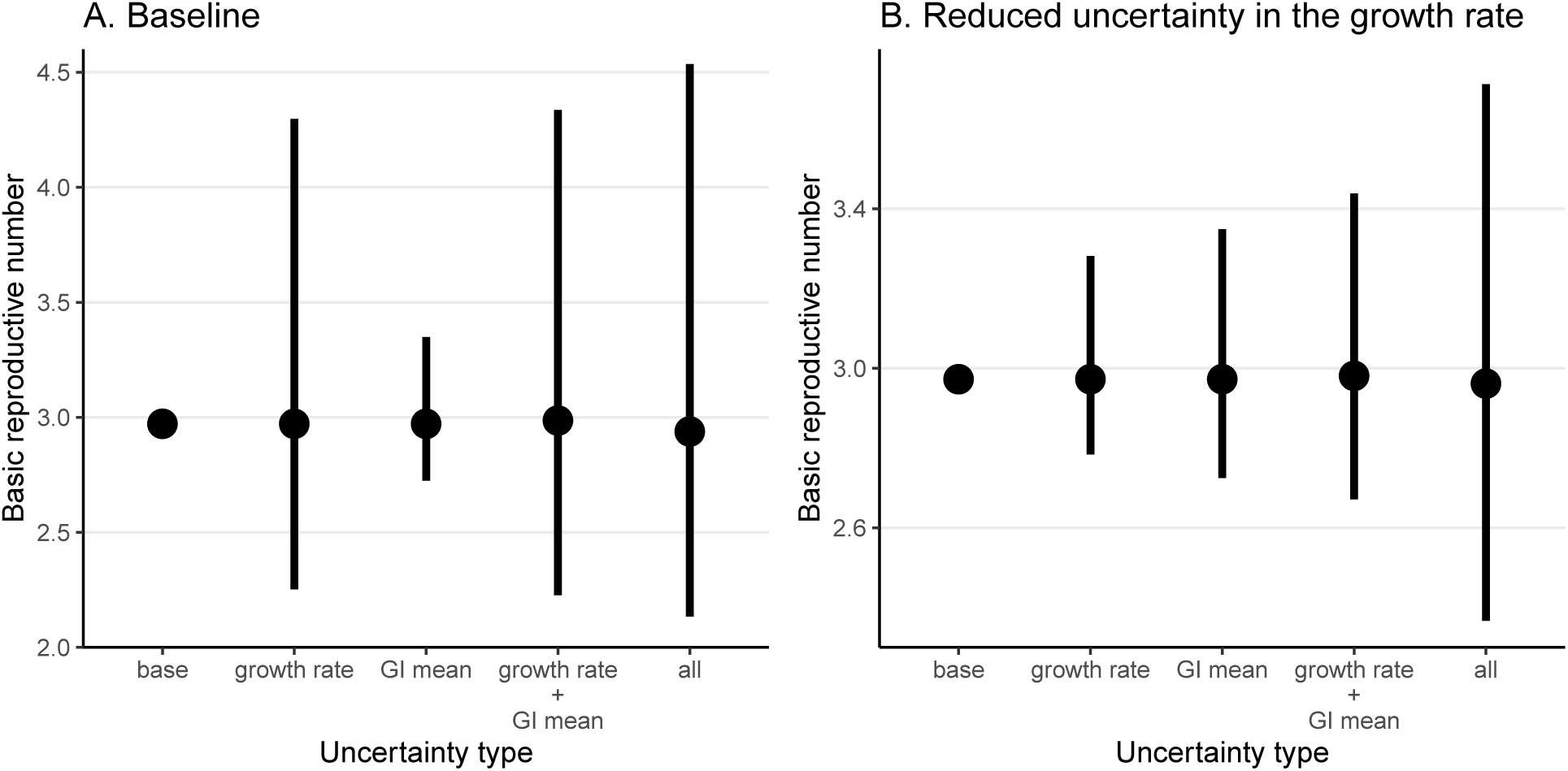
Effects of *r*, 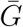, and *κ* on the estimates of *ℛ*_0_. We compare estimates of *ℛ*_0_ under five scenarios that propagate different combinations of uncertainties (A) based on our pooled estimates (*µ*_*r*_, *µ*_*G*_, and *µ*_*κ*_) and (B) assuming a 4-fold reduction in uncertainty of our pooled estimate of the exponential growth rate (using (*µ*_*r*_ + 3 *×* median(*µ*_*r*_))*/*4, instead). **base**: *ℛ*_0_ estimates based on the median estimates of *µ*_*r*_, *µ*_*G*_, and *µ*_*κ*_. **growth rate**: *ℛ*_0_ estimates based on the the posterior distribution of *µ*_*r*_ while using median estimates of *µ*_*G*_ and *µ*_*κ*_. **GI mean**: *ℛ*_0_ estimates based on the the posterior distribution of *µ*_*G*_ while using median estimates of *µ*_*r*_ and *µ*_*κ*_. **growth rate + GI mean**: *ℛ*_0_ estimates based on the the joint posterior distributions of *µ*_*r*_ and *µ*_*G*_ while using a median estimate of *µ*_*κ*_. **all**: *ℛ*_0_ estimates based on the joint posterior distributions of *µ*_*r*_, *µ*_*G*_, and *µ*_*κ*_. Vertical lines represent the 95% confidence intervals.

As uncertainty associated with the exponential growth rate decreases, accounting for uncertainties in generation intervals becomes even more important (Fig. 2B). Propagating error only from the growth rate gives very narrow confidence intervals in this case. Likewise, propagating errors from the growth rate and the mean generation interval gives wider but still too narrow confidence intervals. We expect this hypothetical example to better reflect more recent scenarios, as increased data availability will allow researchers to estimate *r* with more certainty.

We also compare the estimates of *ℛ*_0_ across different studies by replacing their values of *r*, 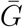, and *κ* with our pooled estimates (*µ*_*r*_, *µ*_*G*_, and *µ*_*κ*_, respectively) one at a time and recalculating the basic reproductive number *ℛ*_0_ (Fig. 3). This procedure allows us to assess the sensitivity of the estimates of *ℛ*_0_ across appropriate ranges of uncertainties. We find that incorporating uncertainties one at a time increases the width of the confidence intervals in all but 7 cases. We estimate narrower confidence intervals for Study 3, Study 6, and Study 7 when we account for proper uncertainties in the generation-interval dispersion because they assume a narrow generation-interval distribution (compare “base” with “GI variation”); when higher values of *κ* are used, their estimates of *ℛ*_0_ become less sensitive to the values of *r* and 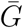, giving narrower confidence intervals. We estimate narrower confidence intervals for Study 5 and Study 7 when we account for proper uncertainties in the mean generation interval (compare “base” with “GI mean”) because the range of uncertainty in the mean generation interval 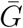 they consider is much wider than the pooled range (Fig. 1). Substituting the reported *r* or 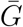 from Study 1 with our pooled estimates give narrower confidence intervals for similar reasons.

**Figure 3:**
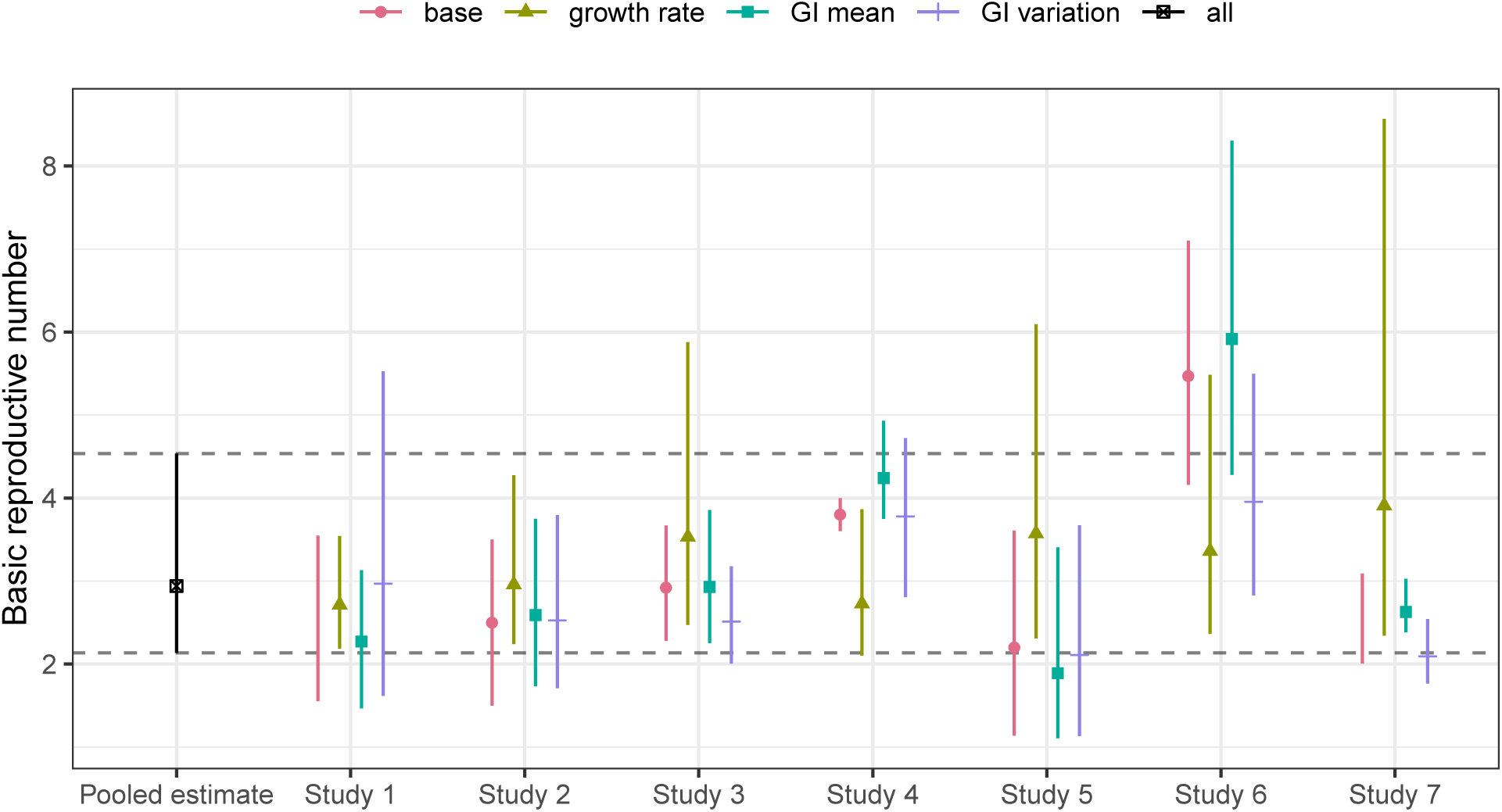
Sensitivity of the reported *ℛ*_0_ estimates with respect to our pooled estimates of the underlying parameters. We replace the reported parameter values (growth rate *r*, GI mean 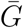, and GI variation *κ*) with our corresponding pooled estimates (*µ*_*r*_, *µ*_*G*_, and *µ*_*κ*_) one at a time and recalculate *ℛ*_0_ (**growth rate, GI mean**, and **GI variation**). The pooled estimate of *ℛ*_0_ is calculated from the joint posterior distribution of *µ*_*r*_, *µ*_*G*_, and *µ*_*κ*_ (**all**); this corresponds to replacing all reported parameter values with our pooled estimates, which gives identical results across all studies. Horizontal dashed lines represent the 95% confidence intervals of our pooled estimate of *ℛ*_0_. The reported *ℛ*_0_ estimates (**base**) have been adjusted to show the approximate 95% confidence interval using the probability distributions that we defined if they had relied on different measures for parameter uncertainties.

We find that accounting for uncertainties in the estimate of *r* has the largest effect on the estimates of *ℛ*_0_ in most cases (Fig. 3). For example, recalculating *ℛ*_0_ for Study 7 by using our pooled estimate of *r* gives *ℛ*_0_ = 3.9 (95% CI: 2.3–8.6), which is much wider than the uncertainty range they reported (2.0–3.1). There are two explanations for this result. First, even though the exponential growth rate *r* and the mean generation interval 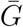 have identical mathematical effects on *ℛ*_0_ in our framework (Eq. 2 in Methods), *r* is more influential in this case because it is associated with more uncertainty (Fig. 1). Second, assuming a fixed generation interval (*κ* = 0) makes the estimate of *ℛ*_0_ too sensitive to *r* and 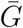. One exception is Study 1: we find this estimate of *ℛ*_0_ is most sensitive to generation-interval dispersion *κ*. This is because Study 1 assumes an exponentially distributed generation interval (*κ* = 1): estimates that rely on this assumption make *ℛ*_0_ relatively insensitive and thus tend to have particularly narrow confidence intervals.

Finally, we incorporate all uncertainties by using posterior samples for *µ*_*r*_, *µ*_*G*_, and *µ*_*κ*_ to recalculate *R*_0_ and compare it with the reported *ℛ*_0_ estimates. Our estimated *ℛ*_0_ from the pooled distribution has a median of 2.9 (95% CI: 2.1–4.5). While the point estimate of *ℛ*_0_ is similar to other reported values from this date range, the confidence intervals are wider than all but one study. This result does not imply that assumptions based on the pooled estimate are too weak; we believe that this confidence interval more accurately reflects the level of uncertainties present in the information that was available when these models were fitted. In fact, because the pooled estimate does not account for overlap in data sources used by the models, we feel that it is more likely to be over-confident than under-confident. Our median estimate averages over the various studies, and therefore particular studies have higher or lower median estimates. We note in particular that, while the baseline example we used from Study 6 may appear to be an outlier, the authors of this study also explore different scenarios involving changes in reporting rate over time, under which their estimates of *ℛ*_0_ are similar to other reported estimates. Here, our focus is on estimating uncertainty, not on identifying potential explanations for these discrepancies.

## 4 Discussion

Estimating the basic reproductive number *ℛ*_0_ is crucial for predicting the course of an outbreak and planning intervention strategies. Here, we use a gamma approximation (Park et al., 2019) to decompose *ℛ*_0_ estimates into three key quantities (*r*, 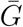, and *κ*) and apply a multilevel Bayesian framework to compare estimates of *ℛ*_0_ for the novel coronavirus outbreak. Our results demonstrate the importance of accounting for uncertainties associated with the underlying generation-interval distributions, including uncertainties in the amount of dispersion in the generation intervals. Our analysis of individual studies shows that many early estimates of *ℛ*_0_ rely on strong assumptions.

Of the seven studies that we reviewed, two of them directly fit their models to cumulative number of confirmed cases. This approach can be appealing because of its simplicity and apparent robustness, but fitting a model to cumulative incidence instead of raw incidence can both bias parameters and give overly narrow confidence intervals, if the resulting non-independent error structures are not taken into account (Ma et al., 2014; King et al., 2015). Naive fits to cumulative incidence data should therefore be avoided.

Many sources of noise affect real-world incidence data, including both dynamical, or “process”, noise (randomness that directly or indirectly affects disease transmission); and observation noise (randomness underlying how many of the true cases are reported). Disease modelers face the choice of incorporating one or both of these in their data-fitting and modeling steps. This is not always a serious problem, particularly if the goal is inferring parameters rather than directly making forecasts (Ma et al., 2014). Modelers should however be aware of the possibility that ignoring one kind of error can give overly narrow confidence intervals (King et al., 2015; Taylor et al., 2016).

There are other important phenomena not covered by our simple framework. Examples that seem relevant to this outbreak include: changing reporting rates, reporting delays (including the effects of weekends and holidays), and changing generation intervals. For emerging pathogens such as SARS-CoV-2, there may be an early period of time when the reporting rate is very low due to limited awareness or diagnostic resources; for example, Zhao et al. (2020) (Study 6) demonstrated that estimates of *ℛ*_0_ can change from 5.47 (95% CI: 4.16–7.10) to 3.30 (95% CI: 2.73–3.96) when they assume 2-fold changes in the reporting rate between January 17, when the official diagnostic guidelines were released (World Health Organization, 2020b), and January 20. Delays between key epidemiological timings (e.g., infection, symptom onset, and detection) can also shift the shape of an observed epidemic curve and, therefore, affect parameter estimates as well as predictions of the course of an outbreak (Tariq et al., 2019). Even though a constant delay between infection and detection may not affect the estimate of the growth rate, it can still affect the associated confidence intervals. Finally, generation intervals can become shorter throughout an epidemic as intervention strategies, such as quarantine, can reduce the infectious period (Hethcote et al., 2002). Accounting for these factors is crucial for making accurate inferences.

Here, we focused on the estimates of *ℛ*_0_ that were published within a very short time frame (January 23–26). During early phases of an outbreak, it is reasonable to assume that the epidemic grows exponentially (Anderson and May, 1991). However, as the number of susceptible individuals decreases, the epidemic will saturate, and estimates of *r* used for *ℛ*_0_ should account for the possibility that *r* is decreasing through time. Although our analysis only reflects a snapshot of a fast-moving epidemic, we expect certain lessons to hold: confidence intervals must combine different sources of uncertainty. In fact, as epidemics progress and more data becomes available, it is likely that inferences about exponential growth rate (and other epidemiological parameters) will become more precise; thus the risk of over-confidence when uncertainty about the generation-interval distribution is neglected will become greater.

We strongly emphasize the value of attention to accurate characterization of the transmission chains via contact tracing and better statistical frameworks for inferring generation-interval distributions from such data (Britton and Scalia Tomba, 2019). A combined effort between public-health workers and modelers in this direction will be crucial for predicting the course of an epidemic and controlling it. We also emphasize the value of transparency from modelers. Model estimates during an outbreak, even in pre-prints, should include code links and complete explanations. We suggest using methods based on open-source tools allow for maximal reproducibility.

In summary, we have provided a basis for comparing exponential-growth based estimates of *ℛ*_0_ and its associated uncertainty in terms of three components: the exponential growth rate, mean generation interval, and generation interval dispersion. We hope this framework will help researchers understand and reconcile disparate estimates of disease transmission early in an epidemic.

## Data Availability

R code is available in GitHub (https://github.com/parksw3/nCoV_framework).

## Acknowledgements

We thank Daihai He for providing helpful comments on the manuscript.

## Contribution

SWP and JD developed the statistical framework. SWP reviewed the published literature. SWP performed the analysis. SWP, BMB, and JD created the figures. SWP and JD wrote the first draft. All authors contributed to the writing and approval of the final report.

## Data availability

R code is available in GitHub (https://github.com/parksw3/nCoV_framework).

**Appendix**

**Figure A1:**
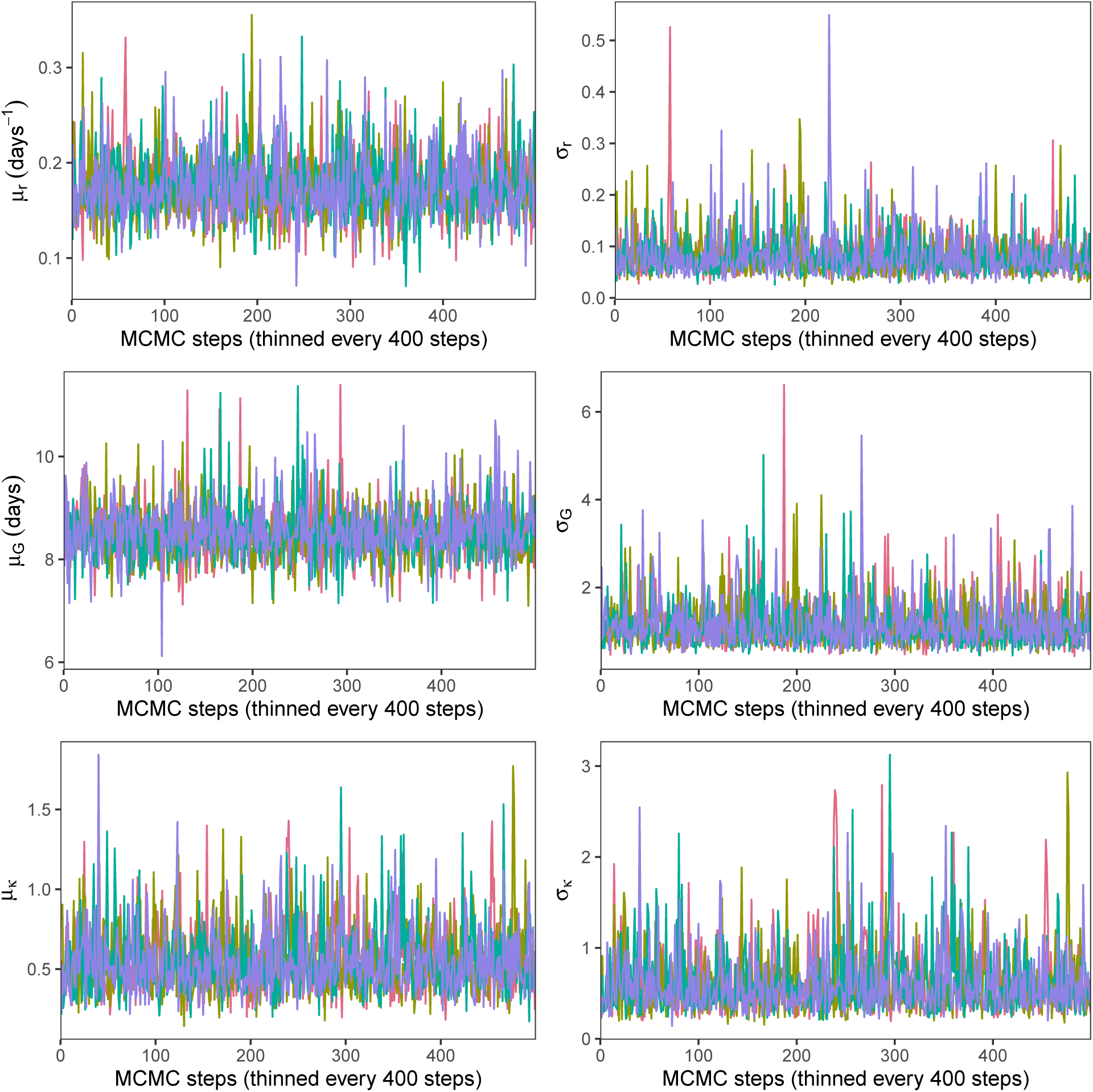
Trace plots of the multilevel model. Each chain is represented by a different color.

**Figure A2:**
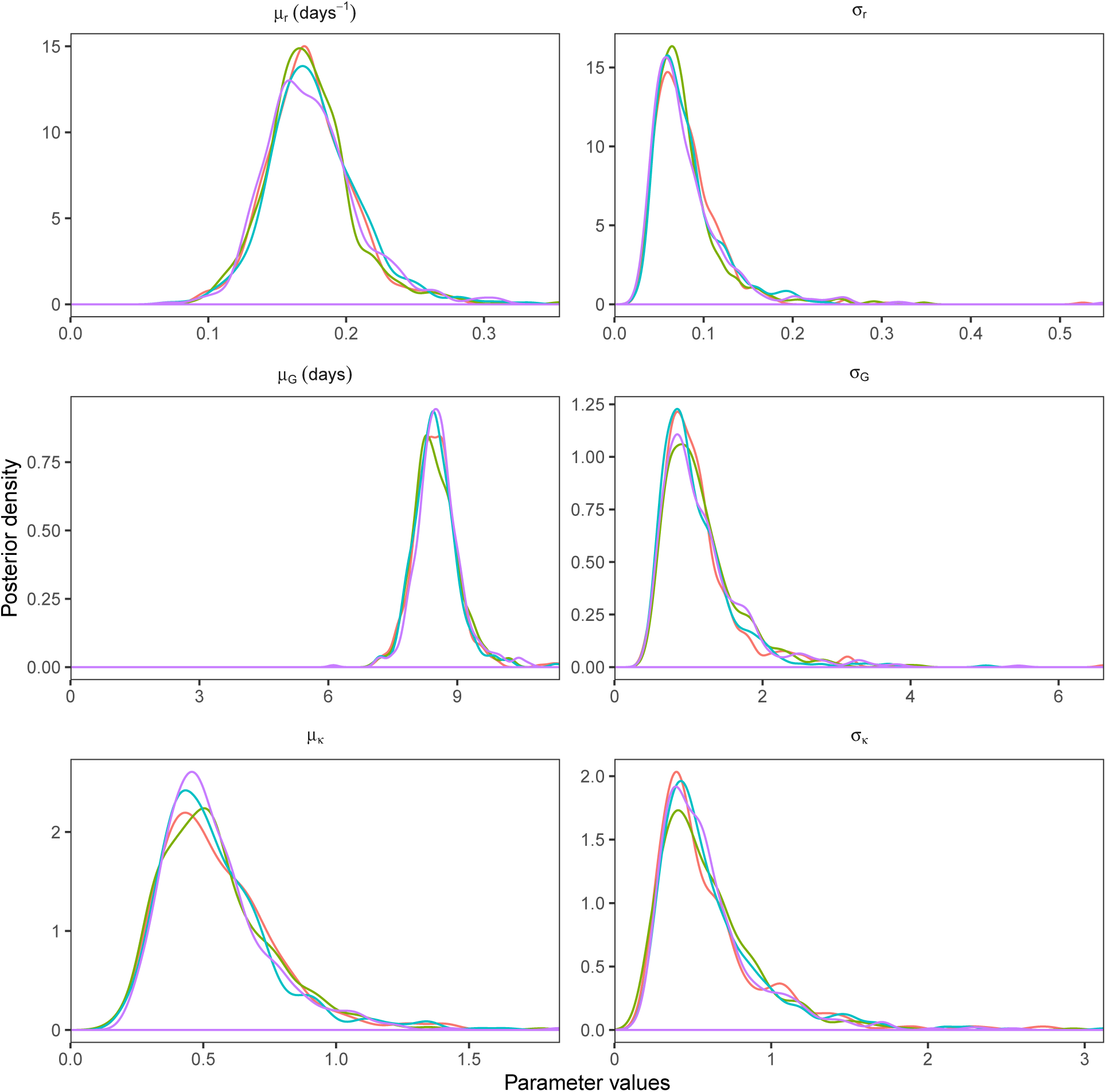
Marginal posterior distributions of the multilevel model. Each chain is represented by a different color.

